# SARS-CoV-2 Seroprevalence in a Cohort of Asymptomatic, RT-PCR Negative Croatian First League Football Players

**DOI:** 10.1101/2020.10.30.20223230

**Authors:** Adriana Vince, Renata Zadro, Zvonimir Šostar, Sunčanica Ljubin Sternak, Jasmina Vraneš, Vedrana Škaro, Petar Projić, Vilim Molnar, Vid Matišić, Bruno Baršić, Gordan Lauc, Zvjezdana Lovrić-Makarić, Zoran Bahtijarević, Tomislav Vlahović, Sandra Šikić, Ozren Polašek, Dragan Primorac

**Author notes:** Correspondence: Adriana Vince, University of Zagreb Medical School, 10000 Zagreb, Mirogojska 8.

## Abstract

**Background:** During the COVID-19 pandemic the Croatian Football Federation has launched a new model of pre-season systematic examination of football players, emphasizing the diagnosis of asymptomatic SARS-CoV-2 infection and preventing further spread among the players.

**Objectives:** The aim of this study was to assess the prevalence and dynamics of SARS-CoV-2 IgA and IgG antibodies in the cohort of asymptomatic and SARS-CoV-2 PCR negative professional football players in the Croatian First Football League by using a commercial ELISA antibody assay in the paired serum samples taken 2 months apart.

**Methods:** Serology testing was performed from May till July 2020 in a cohort of 305 asymptomatic football players and club staff members. RT-PCR for detection of SARS-CoV-2 from nasopharyngeal swabs was performed on three occasions, and Euroimmun ELISA for detection of IgA and IgG (S1 and NCP) antibodies was tested in paired serum samples in May and July.

**Results:** All RT-PCR results were negative. Sixty-one (20%) participants were reactive in one or two classes of antibodies at baseline and/or follow-up serology testing. IgA reactivity was found in 41 (13.4% [95% CI=10.7-17.7]) baseline sera and 42 (13.8% [95% CI=10.3-18.9]) follow-up sera. IgG to S1 protein was found in 6 (2% [95% CI=0.9-4.2]) participants at baseline and 1 (0.33% [95% CI=0.0006-1.83]) at follow-up. IgG to NCP was found in 2 (0.7% [95% CI=0.2-2.4]) participants at baseline and 8 (2.6% (95% CI=1.3-5.1]) participants at follow-up. Noticeable dynamics in the paired sera was observed in 18 (5.9%) participants (excluding borderline IgA results) or 32 (10.5%) (including IgA borderline results).

**Conclusion:** Various patterns of IgA and IgG reactivity were found in the paired serum samples. Based on serology dynamics we estimate that in 5.9%-10.5% of PCR negative football players asymptomatic exposure to SARS-CoV-2 during pandemics could not be excluded.

## Introduction

The COVID-19 outbreak has affected all areas of life including professional sport (1). The pandemic has stopped the sporting calendar, with professional leagues around the globe suspending their activities to limit the virus’s spread and confined the players to an individual training regimen (2). SARS-CoV-2 virus is mainly transmitted via droplets and aerosol generated by sneezing, coughing and talking, as well as through contaminated hands since the virus can survive on various surfaces for several hours or even days (3,4). The diagnosis of COVID-19 includes clinical criteria, epidemiological history and molecular detection of viral RNA in clinical samples. Serological assays that detect specific antibodies to SARS-CoV-2 can be useful in various settings to identify the individuals that remained asymptomatic but should not be used as a standalone test to detect acute infection (5, 6).

The median time to onset of symptoms for persons infected with SARS-CoV-2 is 5 days (7). Studies suggest that the proportion of asymptomatic COVID-19 cases ranges from 17.9% to 78% (8, 9, 10). There is increasing evidence that asymptomatic or minimally symptomatic persons can spread the virus, particularly during the late incubation period, so it is of utmost importance to detect the asymptomatic spreaders timely to prevent rapid disease transmission (11, 12). The immune response in asymptomatic individuals has not been completely understood (13). The football community worldwide is keen to return to football activities, but the return to sporting activities requires risk assessment based on SARS-CoV-2 prevalence within a given cohort and implementation of strict epidemiologic preventive measures accordingly (14). During lock-down in Croatia, the football championship was interrupted as it was the case worldwide. The Croatian Football Federation has launched a new model of pre-season systematic examination of football players, coaches and staff members, with a particular emphasis on diagnosing asymptomatic SARS-CoV-2 infection to prevent further virus spread and to continue safely with training and matches. The model includes epidemiological interviews combined with molecular and serological testing of players before and during the re-starting phase of the first league championship to enable the continuation of the first league games, although without spectators (15).

As repeated molecular testing of SARS-CoV-2 from nasopharyngeal swabs was negative in all tested individuals in our cohort, in this paper we analyzed the serological findings to explore the possibility of asymptomatic exposure to the virus. Earlier studies have found that serum antibodies begin to rise one week after a coronavirus infection with IgA and IgM peaking in the first 5-7 days and decline after 28 days, while IgG antibodies can be detected 7-10 days after infection, reaching the peak 7 weeks later, with long-term memory plasma cells persisting for a long time, protecting individuals against reinfection (16, 17). Seroconversion is faster and more robust in patients with severe disease (6). The duration of detectable antibodies and their neutralizing capacity is still being studied (18, 19). For a more accurate interpretation of the serological result, paired serum specimens from the same individual should be collected at least several weeks apart (6). Various commercial assays utilizing different techniques that measure the binding of IgG, IgM, and/or IgA antibodies have been developed. The performance of the serologic assays varies in different testing cohorts and it has not been fully understood yet (20, 21, 22). The cross-reactivity to other coronaviruses and other viruses can lead to false-positive results (23).

The aim of this study is to analyze the prevalence and dynamics of SARS-CoV-2 IgA and IgG antibodies in the cohort of asymptomatic and SARS-CoV-2 RT-PCR negative professional football players in the Croatian First Football League during the COVID-19 pandemic in Croatia by using a commercial ELISA antibody assay in paired serum samples taken two months apart during COVID-19 pandemic in Croatia.

## Methods

### Study design

Prospective cohort study as a part of Croatian preseason football preparation in the era of COVID-19 performed from April until July of 2020 (15).

### Study Population

A total of 350 participants including all registered football players and club staff members of the Croatian First Football National League participated in this study, all of whom were male aged from 17 to 71 years. All participants were enrolled at the primary club setting. The participants strictly followed the Croatian Football Federation protocol that included limited social contact and training in small groups. None of the participants presented with fever or any respiratory symptoms at the time of testing. An epidemiological questionnaire was filled out at the beginning of the study. One participant was excluded from the study for not completing the questionnaire.

At the time of the third sampling 44 participants were lost to follow-up; therefore, the final number of included participants in the study was 305.

Ethical approval for the study was gained from the Ethics Committee of the Institute of Public Health “dr. Andrija Štampar”. All of the included patients gave a signed written consent to be included in the study prior to testing.

### Sampling

The testing was performed in three phases from May to July 2020. In phase 1 (last week of May 2020), a nasopharyngeal swab was taken for RT-qPCR molecular analysis and the epidemiological questionnaire was filled out by the participants (Supplementary Table 1). In phase 2 (five days after the initial sampling) another nasopharyngeal sample was taken for molecular testing and a sample of peripheral venous blood was drawn for serology. In phase 3 (last week of July), two months after phase 2, both molecular and serological testing was repeated.

**Table 1.** Age and club functions of participants (N=349) by SARS-CoV-2 IgA antibody status.

|  | *IgA negative*<br>(N=301) | *IgA positive*<br>(N=48) | *Total*<br>(N=349) | *p-value* |
| --- | --- | --- | --- | --- |
| *Age* |  |  |  | 0.354 |
| *Mean* | 28.8 | 26.8 | 28.5 |  |
| *SD* | 9.3 | 7.7 | 9.1 |  |
| *Median* | 25 | 24 | 25 |  |
| *25th percentile* | 23 | 22 | 22 |  |
| *75th percentile* | 32 | 29 | 31 |  |
| *Club function* |  |  |  | 0.983 |
| *Player* | 234 (77.7%) | 38 (79.2%) | 272 (77.9%) |  |
| *Coach* | 36 (12.0%) | 7 (14.6%) | 43 (12.3%) |  |
| *Medical staff* | 24 (8.0%) | 2 (4.2%) | 26 (7.4%) |  |
| *Other* | 7 (2.3%) | 1 (2.1%) | 8 (2.3%) |  |

IgA, immunoglobulin A; SD, standard deviation.

In all 3 phases sampling was performed in club ambulances by the same team doctors. Molecular analysis in phases 1 and 2 was performed in the Department of Microbiology of the Institute for Public Health “Dr. Andrija Štampar”, in phase 3 it was performed in Genos Ltd, DNA Laboratory. The serological testing was performed in St. Catherine Specialty Hospital laboratory.

### RT-PCR

Nasopharyngeal swabs were transported in 1.5 mL of Hanks’ Balanced Salt Solution (prepared following in house recipe) at +4 °C to the molecular laboratory.

In phases 1 and 2, isolation of RNA was performed by EZ1 Virus Mini Kit v2.0 (Qiagen, Hilden, Germany on EZ1 Advanced XL instrument for automated purification of nucleic acids (Qiagen, Hilden, Germany). Amplification and detection of SARS-CoV-2 were performed using GeneFinder COVID-19 Plus RealAmp Kit (Osang Healthcare Co. Ltd., Anyang (Dongan), Gyeonggi, South Korea) on Cobas Z 480 Instrument (Roche Diagnostics GmbH, Mannheim, Germany) according to the manufacturer’s protocol. The assay identifies the virus by multiplex real-time RT-PCR targeting three virus genes: the envelope protein (E), the nucleocapsid protein (N) and RNA-dependent RNA polymerase (RdRp) genes. Besides primers for targeted genes, the kit includes the RNase P (RP) primer and probe set for detection of human RP in order to control for specimen quality and demonstration that nucleic acid was generated by the extraction process.

In phase 3 iAMP COVID-19 Detection Kit (ATILA BioSystems, Mountain View, CA, USA) was used for SARS-Cov-2 RNA detection. Raw samples without the RNA extraction process were used. Option C1 - centrifugation method (Recommended validation procedures for iAMP COVID-19 Detection Kit v 2.2, April 2020; protocol provided by the kit manufacturer) was followed for specimen processing and reaction assembly for isothermal amplification. Reverse transcription and amplification of target RNA sequences were performed on MIC device (Bio Molecular Systems, Upper Coomer QLD, Australia), using SARS-CoV-2-specific N/ORF-1ab primer sets.

### Serology

For serologic testing in our cohort, we have decided to use the CE marked commercial ELISA assay (Anti-SARS-CoV-2 IgA, Anti-SARS-CoV-2 IgG S and Anti-SARS-CoV-2 IgG NCP, Euroimmun Medizinische Labordiagnostika AG, Lübeck, Germany) which detects IgA and IgG antibodies to the S1-domain of spike protein, as well as IgG antibodies to nucleocapsid antigen (NCP), which is the antigen with the strongest immune dominance in the coronavirus family. This assay has been validated in numerous studies, showing adequate sensitivity and specificity (24-30). The assay is intended for use as an aid in identifying individuals with an adaptive immune response to SARS CoV-2, indicating recent or prior infection (30). The test was performed on Euroimmun I-2P ELISA analyzer (Euroimmun Medizinische Labordiagnostika AG, Lübeck, Germany) according to the instructions of the manufacturer. After adding the conjugate, a sample’s immunoreactivity was determined by measuring optical density at 450 nm (OD450) and then divided by the OD450 of the calibrator provided to minimize the inter-assay variation. The semiquantitative results were expressed in arbitrary units as OD ratio and interpreted as positive, borderline or negative according to the manufacturer’s proposed cut-off values (≥ 1.1 positive; ≥ 0.8 – < 1.1 borderline; < 0.8 negative). The internal quality control was performed by parallel testing of 6 positive samples from patients who were symptomatic and RT-qPCR confirmed COVID-19, 4-8 weeks before serology testing. The OD ratios from positive controls were: IgA (S1): 0.81-2.58; IgG (S1): 1.65-7.43; IgG (NCP): 3.48-5.07.

To exclude the acute Epstein-Barr virus (EBV) infection and the presence of heterophilic antibodies that could cause cross-reactivity in tested samples, serological analyses were performed to determine IgM and IgG class antibodies against Epstein-Barr virus capsid antigen (Anti-EBV-CA IgM and Anti-EBV-CA IgG, respectively) and IgG against EBV nuclear antigen 1 (Anti-EBNA-1 IgG) supplied by Euroimmun Medizinische Labordiagnostika AG, Lübeck, Germany.

### Statistical analysis

The analysis was performed using SAS 9.4 program, SAS Institute, Cary, North Carolina. Continuous variables were presented as mean, standard deviation, median and 25/75 percentile. Categoric variables were presented as frequencies and percentages. The confidence interval for the proportion of IgA and IgGs was calculated using the Wilson scoring interval (31). We have compared groups of IgA positive and negative individuals. Statistical significance was determined by the Mann-Whitney test for continuous variables and the chi-squared test for categoric variables; for small sample sizes, the exact test was performed. The frequency of positivity in two periods was compared with the McNemar test for dependent samples (Supplementary Table 2).

**Table 2.** SARS-CoV-2 IgA antibody status of participants (N=349) for selected self-reported epidemiological categories.

|  | *IgA negative*<br>(N=301) | *IgA positive*<br>(N=48) | *Total*<br>(N=349) | *p-value* |
| --- | --- | --- | --- | --- |
| *Self-isolation* |  |  |  | 0.985 |
| *Yes* | 11 (3.7%) | 2 (4.2%) | 13 (3.7%) |  |
| *No* | 290 (96.3%) | 46 (95.8%) | 336 (96.3%) |  |
| *Traveling abroad* |  |  |  | 0.845 |
| *Yes* | 19 (6.3%) | 2 (4.2%) | 21 (6.0%) |  |
| *No* | 282 (93.7%) | 46 (95.8%) | 328 (94.0%) |  |
| *Close contact with a COVID-19 positive person* |  |  |  | 0.852 |
| *Yes* | 2 (0.7%) | 0 | 2 (0.6%) |  |
| *No* | 299 (99.3%) | 48 (100%) | 347 (99.4%) |  |

IgA, immunoglobulin A.

## Results

### RT-PCR

In phases 1 and 2, all of the tested participants had negative RT-PCR results for the presence of SARS-CoV-2 in nasopharyngeal samples.

In phase 3, SARS-CoV-2 was not detected for 299 nasopharyngeal samples analyzed with the iAMP COVID-19 Detection Kit. Internal control amplification failure was observed for 6 samples even after the test was repeated. However, the serologic results were negative for those participants in phases 2 and 3, therefore they were considered negative for the presence of SARS-CoV-2.

### Sociodemographics

For the sociodemographic data analysis, we present the data collected from 349 participants included in phases 1 and 2 who answered the epidemiologic questionnaire. Because we have noticed that most of the individuals tested positive for IgA, we decided to show the data according to IgA positivity or negativity. The mean age in our cohort was 28.5 years, median 25 years, 272 (77.9%) of participants were active football players, 43 (12.3%) were coaches and 26 (7.4%) were the medical staff. There were no significant differences in IgA positivity according to age, nor function in the club (Table 1).

There was also no statistical significance in IgA positivity regarding self-reported epidemiologic history related to higher exposure to COVID-19 infection including self-isolation, traveling outside Croatia, or being in close contact with SARS-CoV-2 positive patients (Table 2)

### Serology

For the analysis of serology, we present the data from 305 participants for whom the results of paired serum samples were available. The serological testing results in phases 2 and 3 are presented in Tables 3 and 4 respectively. A total of 61 (20%) out of 305 participants sera were reactive in one or two classes of SARS-CoV-2 antibodies at first and/or follow-up serology testing. The majority of participants (331, 94.8%) had positive IgG antibodies to EBV-VCA and EBNA. Not a single case of acute EBV infection was detected that could cross-react with SARS-CoV-2 antibodies.

**Table 3.**
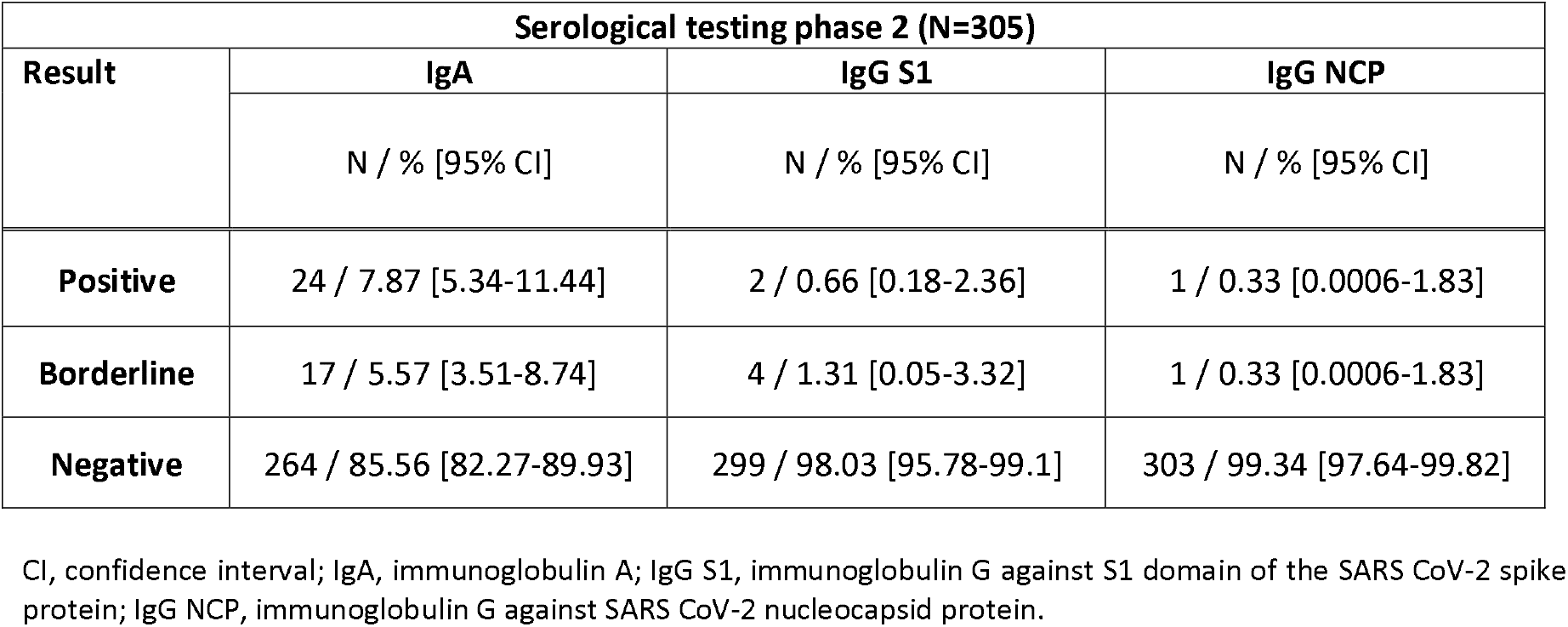
Results of SARS CoV-2 serological testing of participants (N=305) in phase 2.

| Serological testing phase 2 (N=305) |  |  |  |
| --- | --- | --- | --- |
| Result | IgA | IgG S1 | IgG NCP |
|  | N / % [95% CI] | N / % [95% CI] | N / % [95% CI] |
| **Positive** | 24 / 7.87 [5.34-11.44] | 2 / 0.66 [0.18-2.36] | 1 / 0.33 [0.0006-1.83] |
| **Borderline** | 17 / 5.57 [3.51-8.74] | 4 / 1.31 [0.05-3.32] | 1 / 0.33 [0.0006-1.83] |
| **Negative** | 264 / 85.56 [82.27-89.93] | 299 / 98.03 [95.78-99.1] | 303 / 99.34 [97.64-99.82] |

CI, confidence interval; IgA, immunoglobulin A; IgG S1, immunoglobulin G against S1 domain of the SARS CoV-2 spike protein; IgG NCP, immunoglobulin G against SARS CoV-2 nucleocapsid protein.

**Table 4.** Results of SARS CoV-2 serological testing of participants (N=305) in phase 3.

| Serological testing phase 3 (N=305) |  |  |  |
| --- | --- | --- | --- |
| Result | IgA | IgG S1 | IgG NCP |
|  | N / % [95% CI] | N / % [95% CI] | N / % [95% CI] |
| **Positive** | 21 / 6.89 [4.55-10.30] | 0 | 3 / 0.98 [0.34-2.85] |
| **Borderline** | 21 / 6.89 [4.55-10.30] | 1 / 0.33 [0.0006-1.83] | 5 / 3.78 [0.70-1.83] |
| **Negative** | 263 / 86.23 [81.91-89.65] | 304 / 99.67 [98.17-99.94] | 297 / 97.38 [94.91-98.67] |

CI, confidence interval; IgA, immunoglobulin A; IgG S1, immunoglobulin G against S1 domain of the SARS CoV-2 spike protein; IgG NCP, immunoglobulin G against SARS CoV-2 nucleocapsid protein.

The first serology testing (phase 2) showed that 13.4% (95% CI=10.7-17.7) of sera tested borderline or positive for IgA, 2% (95% CI=0.9-4.2) of sera were borderline or positive for IgG (S1) and 0.7% (95% CI=0.2-2.4) were borderline or positive for IgG (NCP). The results were similar at the follow-up testing (phase 3): 13.8% (95% CI=10.3-18.9) borderline or positive results for IgA, only 0.3% (95% CI=0.0006-1.83) IgG (S1) reactive, but 2.6% (95% CI=1.3-5.1) became borderline or positive for IgG (NCP).

The dynamics of the antibody reactivity for each participant is shown in Figure 1.

**Figure 1.**
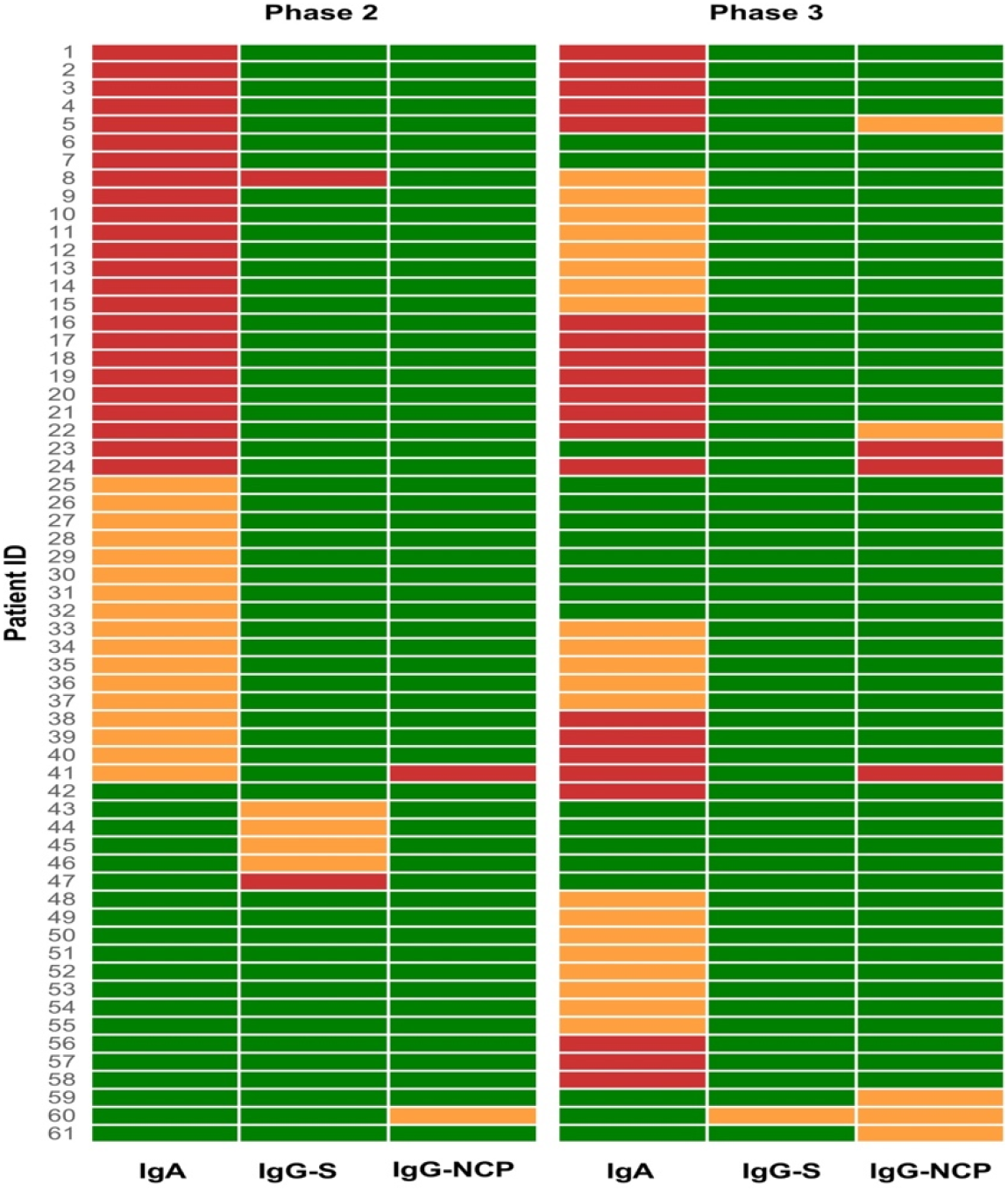
Heatmap of participants serologically positive for SARS-CoV-2 antibodies. Most of the participants were positive in only one class of antibodies. There were only seven cases of double positivity: 2 participants in phase 2: one IgA positive + IgG (S1) positive (participant no. 8) and one IgA borderline + IgG (NCP) positive (participant no 41), 5 participants in phase 3: two IgA positive + IgG (NCP) positive (participants no. 24 and 41), two IgA positive + IgG (NCP) borderline (participants no 5 and 22) and one IgG (S1) borderline + IgG (NCP) borderline (participant no. 60). When we look at the dynamics at the follow-up testing for double-positive sera; the participant no. 8 IgA + IgG (S1) positive stayed borderline in IgA class, but lost IgG antibodies, the participant no. 41 that was IgA borderline and IgG (NCP) positive became IgA and IgG (NCP) positive. Looking at the double positives in the follow-up sera, 3 participants (no. 5, 22 and 24) were already IgA positive at the first serology testing, participant no. 41 was already IgA borderline and IgG (NCP) positive in the first test, and participant no. 60 was IgG (NCP) borderline, so we can conclude that in 6 (2%) out of 305 participants, previous asymptomatic contact with SARS-CoV-2 was fully suspected. Green box, negative; Orange box, borderline; Red box, positive; IgA, immunoglobulin A; IgG S, immunoglobulin G against S1 domain of the SARS CoV-2 spike protein; IgG NCP, immunoglobulin G against SARS CoV-2 nucleocapsid protein.

Participants who were borderline and positive only for the presence of IgA antibodies at baseline were observed with interest at follow-up, because of their relatively high number in the cohort. Thirteen out of 305 (4.2%) participants that were IgA positive (OD ratio >1,1) in the first testing stayed positive in the follow-up, 8/305 (2.6%) IgA positives became IgA borderline at the follow-up, and 3 participants lost their positivity. Out of 17/305 (5.6%) borderline IgA (OD ratio 0.8-1.1), 4/305 (1.3%) became positive (OD ratio >1.1) in the follow-up, 5/305 (1.6%) stayed borderline and 8/305 (2.6%) lost their positivity.

### Assumed SARS-CoV-2 contact

We assumed contact with SARS-CoV-2 when at least 2 types of antibodies were detected at any testing phase and/or when the same class antibody reactivity was found at follow up (mostly IgA). We estimated, according to the dynamics of the antibodies in paired serum samples, that in 5,9% of participants previous contact with SARS-CoV-2 was possible (Table 5). If the borderline results in one of the testing points are regarded as positive, we cannot exclude the previous contact with SARS-CoV-2 in additional 14 (4.6%) participants, therefore a total of 10.5% of participants in our cohort can be suspected of having previous exposure to the virus (Table 5). However, the finding of borderline IgA at both testing points was not assumed as probable contact due to the low specificity of IgA antibodies reported (25, 26, 27).

**Table 5.**
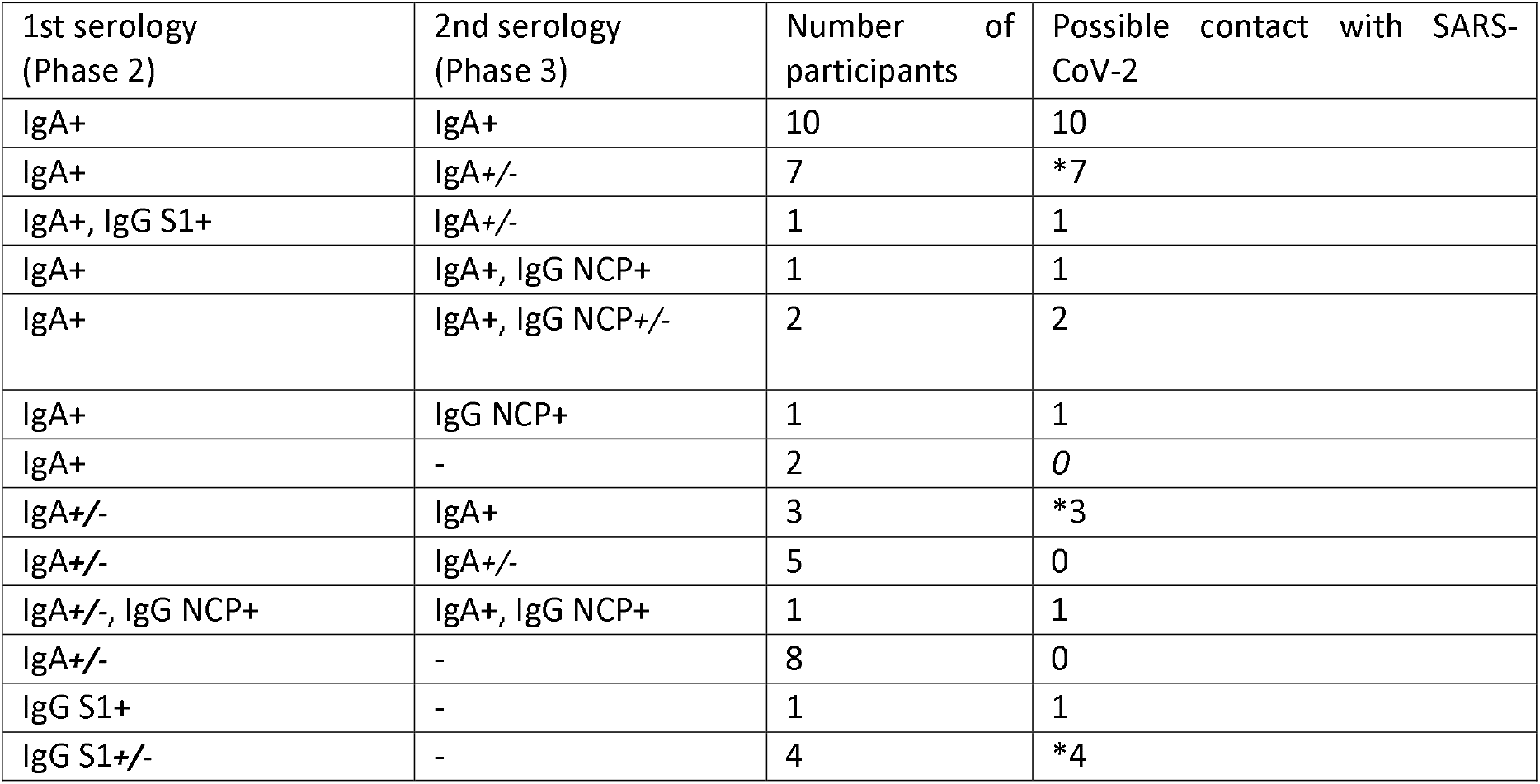
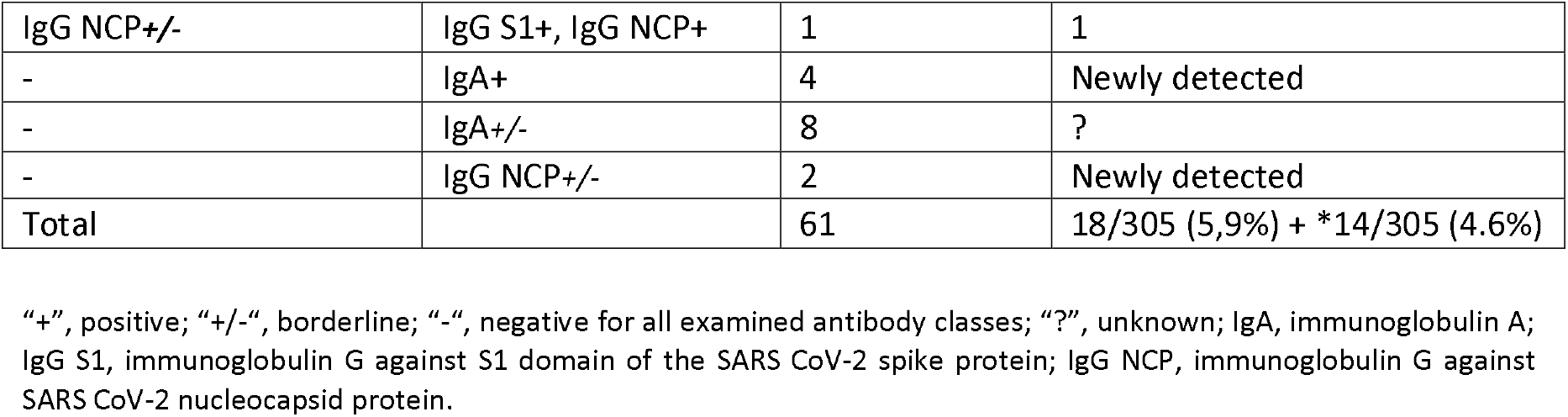
Possible contact with SARS-CoV-2 based on SARS-CoV-2 serology dynamics in paired serum samples of Croatian football players. All of the paired serum combinations are presented. Contact with SARS-CoV-2 is possible in 18 out of 305 participants based on paired serum analysis. If borderline results were interpreted as positive, where only one class of antibodies was borderline in any of the 2 samplings, additional 14 participants could have been in contact with SARS-CoV-2 (as indicated with “*”).

| 1st serology<br>(Phase 2) | 2nd serology<br>(Phase 3) | Number of<br>participants | Possible contact with SARS-CoV-2 |
| --- | --- | --- | --- |
| IgA+ | IgA+ | 10 | 10 |
| IgA+ | IgA+/- | 7 | *7 |
| IgA+, IgG S1+ | IgA+/- | 1 | 1 |
| IgA+ | IgA+, IgG NCP+ | 1 | 1 |
| IgA+ | IgA+, IgG NCP+/- | 2 | 2 |
| IgA+ | IgG NCP+ | 1 | 1 |
| IgA+ | - | 2 | 0 |
| IgA+/- | IgA+ | 3 | *3 |
| IgA+/- | IgA+/- | 5 | 0 |
| IgA+/-, IgG NCP+ | IgA+, IgG NCP+ | 1 | 1 |
| IgA+/- | - | 8 | 0 |
| IgG S1+ | - | 1 | 1 |
| IgG S1+/- | - | 4 | *4 |
| IgG NCP+/- | IgG S1+, IgG NCP+ | 1 | 1 |
| - | IgA+ | 4 | Newly detected |
| - | IgA+/- | 8 | ? |
| - | IgG NCP+/- | 2 | Newly detected |
| Total |  | 61 | 18/305 (5,9%) + *14/305 (4.6%) |

“+”, positive; “+/-”, borderline; “-”, negative for all examined antibody classes; “?”, unknown; IgA, immunoglobulin A; IgG S1, immunoglobulin G against S1 domain of the SARS CoV-2 spike protein; IgG NCP, immunoglobulin G against SARS CoV-2 nucleocapsid protein.

## Discussion

### Study background

SARS-CoV-2 infection can have various clinical presentations: from asymptomatic to severe cases with pneumonitis, ARDS, and multiple organ failure; and possible long-term health consequences, primarily lung fibrosis (32, 33). In young people, most of the infections show a mild or asymptomatic course, however, severe cases with heart complications have also been described (34). Still, those patients can spread the virus to their family, friends and colleagues, especially in the last days of incubation when the quantity of virus is the highest in the respiratory specimens (6). This is important for football players as they have many close contacts during training, traveling, matches, etc. and can quickly spread the virus among their team, coaches, staff and others. Football player’s career development requires a lot of time and effort invested into the prevention of injury, disease, disability and even death, therefore it is crucial to identify the infection early and isolate the infected person from the rest of the team in a timely manner. In this prospective study, we wanted to explore what information on exposition to SARS-CoV-2 we can obtain from serological data in the cohort of first league football players and staff members that were preparing for the restart of football season during the COVID-19 outbreak in Croatia. During the follow-up period from May to July 2020 study participants were tested three times for SARS-CoV-2 by RT-PCR. Not a single case of active infection was detected. All of the participants denied having any respiratory symptoms or fever throughout the whole study period. The seroprevalence of SARS-CoV-2 in the Croatian general population has not yet been published, but similar studies in cohorts of healthcare professionals and factory workers demonstrated that 1.27% and 2.7% tested individuals were positive for the presence of antibodies respectively (35, 36).

### Results comparison

Altogether in 61 out of 305 participants serum reactivity in at least one antibody class was found. We detected IgA positivity in 7.9% and 6.9% at baseline and follow-up sera, respectively. The prevalence of borderline IgA was 5.6% and 6.9% respectively. Euroimmun IgA ELISA was already validated in different studies. Jääskeläinen and coworkers, while evaluating IgG and IgA Euroimmun ELISA test, found lower specificity of IgA of 73% for presumably negative patients, however, the number of screened serum samples was small without follow-up testing. Positive IgA was also detected earlier and more frequently than IgG in serum samples from confirmed COVID-19 patients. The authors conclude that a second convalescent serum is needed to obtain reliable results (25). We have found that IgA remained positive (OD>1.1) in 13 participants in the second testing, so we cannot ignore those results. Nicol et al. reported 17.3% false IgA positives among RT-PCR negatives, but they had tested only 50 presumably negative serum samples on one occasion and the “grey zone” was considered positive (26). In our study, we have found 5.6% IgA borderline results at the first testing and 6.9% at the second testing. Van Elslande et al. found very low ELISA IgA specificity of 73.8% in negative controls and pointed out that the ELISA IgA should not be used for the screening of asymptomatic persons (27). Beavis et al. found 88.4% IgA specificity for 86 negative samples with borderline samples included in the positive results and concluded that results from antibody testing should not be used as the sole basis to diagnose or exclude SARS-CoV-2 infection or to inform infection status (24). Obviously, there is a limitation of ELISA IgA performance due to false-positivity in individuals without symptoms, most probably caused by cross-reactivity to other human coronaviruses.

However, all of the aforementioned studies have investigated only single serum samples from presumably negative individuals or COVID-19 patients, so no dynamics in antibody response could be observed. On the contrary, we have analyzed paired sera and found substantial positive IgA dynamics in 15 out of 305 (5%) of participants (Table 5, Figure 1). Borderline IgA results in asymptomatic patients are not easy to interpret, therefore we decided to leave out of conclusion the participants that had borderline results without any dynamics or presence of IgG antibodies simultaneously. IgG (S1) antibodies were detected in 6 cases only. Meyer and coworkers suggest that higher IgG (S1) cut-off value for seropositivity is needed (2.5) to secure an optimal specificity and positive predictive value (28). However, IgG (S1) was detectable at baseline only in 6 sera in our cohort, out of which 4 were borderline and 2 had positive OD ratios (1.37 and 4.25). IgG (S1) positivity was not observed in paired sera at follow up 2 months later. Ortho-Heller and coworkers found a stronger decrease for IgG (NCP) than for S1 specific antibodies looking at the longitudinal kinetics in the cohort of 20 non-hospitalized patients. They conclude that a single SARS-COV-2 antibody test should not be used to exclude or confirm a previous infection (37). In our cohort IgG (NCP) antibodies were detectable only in two participants in phase 2, which also persisted through phase 3, while 6 new positive participants appeared, indicating possible asymptomatic infection between the two testing points. This is in line with Van Eslandie’s retrospective study, which found a shorter time to seropositivity for IgG (NCP) compared with IgG (S1), with similar specificity for pre-COVID samples 94.7% and 96.55, respectively (38).

### Limitations and added value

This study’s limitations include relatively small sample size, possible cross-reactivity with antibodies to other human coronaviruses that were not analyzed, and the fact that only one commercial ELISA test was used. Also, we were not able to include serum samples from the pre-COVID era. The main added value of the study is the analysis of paired sera tested for IgA and IgG S and NCP antibodies in a homogenous cohort of 305 healthy young male participants, all negative for the presence of SARS-CoV-2 by RT-PCR in 3 testing phases and asymptomatic during the observation period.

## Conclusion

Various patterns of IgA and IgG antibody reactivity were found in the cohort of 305 asymptomatic, RT-PCR negative Croatian first league football players in the paired serum samples collected in the period from May to July 2020. IgA reactivity was predominant and was found in 13.4% of tested sera at baseline and 13.8% at follow-up, either as borderline or positive following the manufacturer’s proposed OD ratios in the ELISA test. According to already published data on false-positive IgA results in presumably negative serum samples, our results should be interpreted cautiously. IgG reactivity was scarce (0.3-2.6%) at both testing points. Based on serology dynamics, we can conclude that in 5.9%-10.5% of PCR negative football players asymptomatic exposure to SARS-CoV-2 during pandemics could not be excluded. This calls for more frequent testing in asymptomatic players, perhaps with rapid antigen tests as point-of-care diagnostics for SARS-CoV-2 (39, 40). It is obvious that SARS-CoV-2 can be transmitted asymptomatically in the cohort of football players, and authorities should insist on the strict implementation of preventive measures during overall sports activities.

## Data Availability

Data are available upon reasonable request.

## Acknowledgments

We thank the Croatian Football Federation and its president Mr. Davor Šuker, for their all-around support in the study. We also thank the Croatian first football league players and staff members for their participation and support in the sample obtainment process.

## References

1 Corsini A, Bisciotti GN, Eirale C, Volpi P. Football cannot restart soon during the COVID-19 emergency! A critical perspective from the Italian experience and a call for action. Br J Sports Med. 2020;54:1186–1187. doi: 10.1136/bjsports-2020-102306.

2 Eirale C, Bisciotti G, Corsini A, Baudot C, Saillant G, Chalabi H. Medical recommendations for home-confined footballers’ training during the COVID-19 pandemic: from evidence to practical application. Biol Sport. 2020;37:203–207. doi: 10.5114/biolsport.2020.94348.

3 van Doremalen N, Bushmaker T, Morris DH, Holbrook MG, Gamble A, Williamson BN et al. Aerosol and Surface Stability of SARS-CoV-2 as Compared with SARS-CoV-1. N Engl J Med. 2020;382:1564–1567. doi: 10.1056/NEJMc2004973.

4 Kampf G, Todt D, Pfaender S, Steinmann E. Persistence of coronaviruses on inanimate surfaces and their inactivation with biocidal agents. J Hosp Infect. 2020;104:246–251. doi: 10.1016/j.jhin.2020.01.022.

5 He F, Deng Y, Li W. Coronavirus disease 2019: What we know? J Med Virol. 2020;92:719–725. doi: 10.1002/jmv.25766.

6 WHO. Diagnostic testing for SARS-CoV-2, Interim Guidance, 11 September 2020. Available at: https://www.who.int/emergencies/diseases/novel-coronavirus-2019/technical-guidance-publications

7 Lauer SA, Grantz KH, Bi Q, Jones FK, Zheng Q, Meredith HR et al. The incubation period of coronavirus Disease 2019 (COVID-19) from publicly reported confirmed cases: Estimation and application. Ann Intern Med. 2020;172:577–582. doi: 10.7326/M20-0504.

8 Mizumoto K, Kagaya K, Zarebski A, Chowell G. Estimating the asymptomatic proportion of coronavirus disease 2019 (COVID-19) cases on board the Diamond Princess cruise ship, Yokohama, Japan, 2020. Euro Surveill. 2020;25:2000180. doi:10.2807/1560-7917.ES.2020.25.10.2000180.

9 Day M. Covid-19: four fifths of cases are asymptomatic, China figures indicate. BMJ. 2020;369:m1375. doi: https://doi.org/10.1136/bmj.m1375.

10 Nishiura H, Kobayashi T, Miyama T, Suzuki A, Jung SM, Hayashi K et al. Estimation of the asymptomatic ratio of novel coronavirus infections (COVID-19). Int J Infect Dis. 2020;94:154–155. doi: 10.1016/j.ijid.2020.03.020.

11 Ye F, Xu S, Rong Z, Xu R, Liu X, Deng P et al. Delivery of infection from asymptomatic carriers of COVID-19 in a familial cluster. Int J Infect Dis. 2020;94:133–138. doi: 10.1016/j.ijid.2020.03.042.

12 Rothe C, Schunk M, Sothmann P, Bretzel G, Froeschl G, Wallrauch C, et al. Transmission of 2019-nCoV Infection from an Asymptomatic Contact in Germany. N Engl J Med. 2020;382:970– 971. doi: 10.1056/NEJMc2001468.

13 Long Q, Tang X, Shi Q, Li Q, Deng H, Yuan J et al. Clinical and immunological assessment of asymptomatic SARS-CoV-2 infections. Nat Med. 2020;26:1200–1204. doi: 10.1038/s41591-020-0965-6.

14 Carmody S, Murray A, Borodina M, Gouttebarge V, Massey A. When can professional sport recommence safely during the COVID-19 pandemic? Risk assessment and factors to consider. Br J Sports Med. 2020;54:946–948. doi: 10.1136/bjsports-2020-102539.

15 Primorac D, Matišić V, Molnar V, Bahtijarević Z, Polašek O. Pre-season football preparation in the era of COVID-19: Croatian Football Association Model. JoGH. 2020;10:010352. doi: 10.7189/jogh.10.010352.

16 Wu L-P, Wang N-C, Chang Y-H, Tian X-Y, Na D-Y, Zhang L-Y et al. Duration of antibody responses after severe acute respiratory syndrome. Emerg Infect Dis. 2007;13:1562–4. doi: 10.3201/eid1310.070576.

17 Stephens DS, McElrath MJ. COVID-19 and the path to immunity. JAMA. 2020;324:1279–1281. doi:10.1001/jama.2020.16656.

18 Zhao J, Yuan Q, Wang H, Liu W, Liao X, Su Y et al. Antibody responses to SARS-CoV-2 in patients of novel coronavirus disease 2019. Clin Infect Dis. 2020:ciaa344. doi:10.1093/cid/ciaa344. [published online ahead of print].

19 Okba N, Müller MA, Li W, Wang C, GeurtsvanKessel CH, Corman VM, et al. Severe Acute Respiratory Syndrome Coronavirus 2−Specific Antibody Responses in Coronavirus Disease Patients. Emerg Infect Dis. 2020;26:1478–1488. doi:10.3201/eid2607.200841.

20 Lisboa Bastos M, Tavaziva G, Abidi SK, Campbell JC, Haraoui LP, Johnston JC et al. Diagnostic accuracy of serological tests for COVID-19: systematic review and meta-analysis. BMJ. 2020;370:m2516. doi: https://doi.org/10.1136/bmj.m2516.

21 Gorse GJ, Donovan MM, Patel GB. Antibodies to coronaviruses are higher in older compared with young adults and binding antibodies are more sensitive than neutralizing antibodies in identifying corona-associated illness. J med Virol. 2020;92:512–517. doi: 10.1002/jmv.25715.

22 GeurtsvanKessel CH, Okba NMA, Igloi Z, Bogers S, Embregts CWE, Laksono BM et al. An evaluation of COVID-19 serological assays informs future diagnostics and exposure assessment. Nat Commun. 2020;11:3436. doi: 10.1038/s41467-020-17317-y.

23 Che XY, Qiu LW, Liao ZY, Wang YD, Wen K, Pan YX et al. Antigenic cross-reactivity between severe acute respiratory syndrome-associated coronavirus and human coronaviruses 229E and OC43. J Infect Dis. 2005;191:2033–2037. doi: 10.1086/430355.

24 Beavis KG, Matushek SM, Abeleda APF, Bethel C, Hunt C, Gillen S et al. Evaluation of the EUROIMMUN Anti-SARS-CoV-2 ELISA Assay for detection of IgA and IgG antibodies. J Clin Virol. 2020;129:104468. doi: 10.1016/j.jcv.2020.104468.

25 Jääskeläinen AJ, Kekäläinen E, Kallio-Kokko H, Mannonen L, Kortela E, Vapalahti O et al. Evaluation of commercial and automated SARS-CoV-2 IgG and IgA ELISAs using coronavirus disease (COVID-19) patient samples. Euro Surveill. 2020;25:2000603. doi:10.2807/1560-7917.ES.2020.25.18.2000603.

26 Nicol T, Lefeuvre C, Serri O, Pivert A, Joubaud F, Dubee V et al. Assessment of SARS-CoV-2 serological tests for the diagnosis of COVID-19 through the evaluation of three immunoassays: Two automated immunoassays (Euroimmun and Abbott) and one rapid lateral flow immunoassay (NG Biotech). J Clin Virol. 2020;129:104511. doi:10.1016/j.jcv.2020.104511.

27 Van Elslande J, Houben E, Depypere M, Brackeneir A, Desmet S, Andre E et al. Diagnostic performance of seven rapid IgG/IgM antibody tests and the Euroimmun IgA/IgG ELISA in COVID- 19 patients. Clin Microbiol Infect. 2020;26:1082–1087. doi:10.1016/j.cmi.2020.05.023.

28 Meyer B, Torriani G, Yerly S, Mazza L, Calame A, Arm-Vernez I et al. Validation of a commercially available SARS-CoV-2 serological immunoassay. Clin Microbiol Infect. 2020;26:1386–1394. doi:10.1016/j.cmi.2020.06.024.

29 Snoeck CJ, Vaillant M, Abdelrahman T, Satagopam VP, Turner JD, Beaumont K et al. Prevalence of SARS-CoV-2 infection in the Luxembourgish population: the CON-VINCE study. MedRxiv. 2020.05.11.20092916. doi: 10.1101/2020.05.11.20092916.

30 FDA:Anti-SARS-VoV-2 ELISA (IgG) Instruction for use. Emergency use authorization. Available at: https://www.fda.gov/medical-devices/emergency-situations-medical-devices-emergency-use-authorization#covid9ivd.

31 Brown DL, Cai TT, DasGupta A. Interval Estimation for a Binomial Proportion. Statis Sci. 2001;16 (2):101–117.

32 Wu D, Shu T, Yang X, Song JX, Zhang M, Yao C et al. Plasma Metabolomic and Lipidomic Alterations Associated with COVID-19. Natl Sci Rev. 2020;nwaa086. doi:10.1093/nsr/nwaa086. [published online ahead of print].

33 Su H, Yang M, Wan C, Yi LX, Tang F, Zhu HY et al. Renal histopathological analysis of 26 postmortem findings of patients with COVID-19 in China. Kidney Int. 2020;98:219–227. doi: 10.1016/j.kint.2020.04.003.

34 Puntmann VO, Carerj ML, Wieters I, Fahim M, Arendt C, Hoffmann J et al. Outcomes of Cardiovascular Magnetic Resonance Imaging in Patients Recently Recovered From Coronavirus Disease 2019 (COVID-19). JAMA Cardiol. 2020:e203557. doi:10.1001/jamacardio.2020.3557. [published online ahead of print].

35 Vilibic-Cavlek T, Stevanovic V, Tabain I, Betica-Radic LJ, Sabadi D, Peric LJ et al. Severe acute respiratory syndrome coronavirus 2 seroprevalence among personnel in the healthcare facilities of Croatia, 2020. Rev Soc Bras Med Trop. 2020;53:e20200458. doi:10.1590/0037-8682-0458-2020.

36 Jerković I, Ljubić T, Bašić Ž, Kružić I, Kunac N, Bezić J et al. SARS-CoV-2 Antibody Seroprevalence in Industry Workers in Split-Dalmatia and Šibenik-Knin County, Croatia. J Occup Environ Med. 2020. doi: 10.1097/JOM.0000000000002020. [published online ahead of print].

37 Orth-Höller D, Eigentler A, Stiasny K, Weseslindtner L, Möst J. Kinetics of SARS-CoV-2 specific antibodies (IgM, IgA, IgG) in non-hospitalized patients four months following infection. J Infect. 2020:S0163-4453(20)30593-4. doi: 10.1016/j.jinf.2020.09.015. [published online ahead of print].

38 Van Elslande J, Decru B, Jonckheere S, Van Wijngaerden E, Houben E, Vandecandelaere P et al. Antibody response against SARS-CoV-2 spike protein and nucleoprotein evaluated by four automated immunoassays and three ELISAs. Clinical Microbiology and Infection 2020:S1198-743X(20)30446-8. doi:10.1016/j.cmi.2020.07.038. [published online ahead of print].

39 WHO. Antigen-detection in the diagnosis of SARS-CoV-2 infection using rapid immunoassays. 11 September 2020. Available at: https://www.who.int/publications/i/item/antigen-detection-in-the-diagnosis-of-sars-cov-2infection-using-rapid-immunoassays.

40 CDC. Interim Guidance for Rapid Antigen Testing for SARS-CoV-2. 4 September 2020.Available at: https://www.cdc.gov/coronavirus/2019-ncov/lab/resources/antigen-testsguidelines.

